# A Cross-Sectional Analysis of Oral Health Spending Over the Lifespan in Commercial and Medicaid Insured Populations

**DOI:** 10.1101/2021.05.17.21256858

**Authors:** Eric P Tranby, Julie Frantsve-Hawley, Myechia Minter-Jordan, James Thommes, Matt Jacob, Michael Monopoli, Ilya Okunev, Sean G. Boynes

**Author notes:** **Corresponding Author:** Julie Frantsve-Hawley, 465 Medford Street, Boston, MA 02129.

## Abstract

**Background:** Life course theory creates a better framework to understand how oral health needs and challenges align with specific phases of the lifespan, care models, social programs, and changes in policy.

**Methods:** Data are from the 2018 IBM Watson Multi-State Medicaid Marketscan Database (31 million claims) and the 2018 IBM Watson Dental Commercial and Medicare Supplemental Claims Database (45 million claims). Analysis compares per enrollee spending fee-for-service dental claims and medical spending on dental care from ages 0 to 89.

**Results:** Dental utilization and spending are lower during the first 4 years of life and in young adulthood than in other periods of life. Stark differences in the timing, impact, and severity of caries, periodontal disease, and oral cancer are seen between those enrolled in Medicaid and commercial dental plans. Early childhood caries and oral cancer occur more frequently and at younger ages in Medicaid populations.

**Conclusions:** This unique lifespan analysis of the U.S. multi-payer dental care system demonstrates the complexities of the current dental service environment and a lack of equitable access to oral healthcare.

**Practical Implications:** Health policies should be focused on optimizing care delivery to provide effective preventive care at specific stages of the lifespan.

---

Because of the life-long process needed to keep the mouth healthy, it is vital to understand how the timing of various structural and behavioral factors—such as historical exposure to disease risk, care administration, socioeconomic impacts, and individual decision making—impact oral health and quality of life. Therefore, models have been developed and proposed to explain how intrinsic and extrinsic events at various periods throughout one’s life can affect oral health.^1–4^ Several of the structural and behavioral factors provided in these models are related to oral health care services utilization and dental care spending.^4–9^

In the U.S., benefit or insurance designs that heavily influence utilization and spending are dichotomized into Commercial and Governmental systems. The benefits offered by Commercial and Medicaid dental insurance are very different. Commercial dental plans generally offer a comprehensive mix of dental services that cover diagnostic, preventive, restorative, periodontal, and sometimes orthodontics/prosthodontic procedures, although most plans are subject to annual and lifetime spending limits along with deductibles or co-pays.^10^ Comprehensive dental coverage is mandatory for children enrolled in Medicaid, but states can choose whether or not to cover dental services for adults in their Medicaid programs.^11^ Coverage varies across the states, and is dependent on state budgets, political dynamics, and public health infrastructure. Even with benefits, barriers to access to care may remain because dental practice operational overhead limits participation in Medicaid plans and many choose not to serve Medicaid enrolled patients.^12^ These coverage variations, along with broader social determinants of health, have wide-ranging impacts including poorer oral health and medical health, and negative influences on employment.^13^ Therefore, understanding the cost and structure of care over a lifespan can allow for a comparison to other systems affecting health, explore how changes in psychosocial structures affect utilization and care delivery costs, and provide continued exploration of how oral health clinical environments impact quality of life.

Gaining a deeper understanding of these relationships could be beneficial by providing an important context for decisions that shape clinical practice, care integration strategies, oral health policy and resource allocation. For these and other reasons, researchers see life-course theory and the accumulation-of-risk model as vehicles for addressing dynamics that shape or are shaped by oral disease.^14^ Life Course theory is a conceptual and theoretical orientation that explains individuals’ health and disease patterns using several key concepts, including lifespan development, human agency, timing, linked lives, and historical time and place.^15^ The concept of lifespan development argues that health and disease patterns are sequential and cumulative, with experiences earlier in life influencing what happens in later life. Life course theory also argues that lives, and health outcomes, are linked based on common shared experiences related to social determinants of health, access to care, clinical treatment patterns, health policy, disease states, and shared historical time and place.^15^ Life-course theory has bi-directional implications, both in how social determinants, such as poverty, nutrition, housing and transportation affect oral health,^16^ and in how oral disease can undermine people’s overall health, psychosocial and economic status. Producing and tracking healthy outcomes over the lifetime has become more important due to: continued changes in reimbursement structure,^17^ improvement in medical-dental care integration,^18–19^ the growth of safety net dental and oral health care models, ^20–22^ improved access to data,^23^ changing patient expectations,^24^ and increased awareness of oral health as an operational concept.^25–27^ In fact, the U.S. Surgeon General announced that lifespan analysis will be a significant component of the upcoming Surgeon General’s Oral Health Report, 2020.^28^

In this article, we employ a cross-sectional lifespan analysis, breaking out spending on dental care and medical spending on oral health by category across each year of life (ages 0-89). The analysis includes claims information from both Medicaid insured and commercially insured patients. This analysis allows a unique perspective into the pattern of spending on dental and oral health care at different stages of the lifespan and emphasizes the role of insurance coverage and policy environment as a key differentiator in the timing, impact, and severity of oral diseases. Employing a lifespan analysis of oral health services and utilization contributes to life course health strategies and can allow stakeholders to better understand how new care models, social programs, grant spending, and changes in policy environments impact society or improve quality of life.

## Methodology

This evaluation provides the results of a cross-sectional analysis, breaking out spending on dental care and medical spending on oral health by category across each year of life (ages 0-89). The data sources for this analysis are the 2018 IBM Watson Multi-State Medicaid Marketscan Database, which represents all Medicaid claims in 13 states during that year, and the 2018 IBM Watson Dental Commercial and Medicare Supplemental Claims Database, which includes all dental claims and medical claims from a convenience sample of IBM Watson data contributors.^29^ This includes 31 million Medicaid dental claims (covering 5.7 million enrollees and 1.1 million patients) and over 45 million dental claims (covering 6.6 million enrollees and 4 million patients) from commercial insurance or Medicare supplemental plans over a lifespan. Importantly, these data also include information on medical utilization for dental conditions, allowing us to estimate the burden on the medical system of treating oral health diseases.

This analysis is restricted to enrollees in fee-for-service reimbursement systems, as indicated by their last period of enrollment during the year. The commercial database is further restricted to those covered by both dental and medical insurance. While dental and medical claims among patients eligible for Medicare are included in both databases, claims paid by Medicare are not included in the Medicaid database; it is likely that some claims are missing for those patients, especially those dual-eligible for Medicaid and Medicare.

Cost is assessed using a variable that reflects the amount paid to the provider for the service. This variable reflects both payments to the provider from a third-party insurer and direct payments to the provider from the patient in co-pays or co-insurance amounts. Average costs per enrollee are calculated to normalize costs across the population eligible for services. Without such normalization, it would be impossible to compare costs associated with medical spending on non-traumatic dental conditions with dental spending, due to vast differences in utilization as well as the prevalence of conditions like oral cancer. Alternative ways of calculating and normalizing costs for dental patients are considered in supplemental analyses. Dental spending is split into categories to better understand how costs change over the lifetime. Spending on dental care is restricted to procedures performed by dentists. Categories, which utilize the American Dental Association’s Code on Dental Procedures and Nomenclature (CDT) 2018, were selected to ease comparison of the large data sets: Preventive care and basic procedures, a category comprised of procedures related to diagnostics, prevention, and less invasive intervention; and major procedures, a category comprised of procedures related to disease stabilization, which are often irreversible, and more invasive interventions.^30^ Preventive care and basic procedures include diagnostic, imaging, preventive, and minor restoration. Major dental procedures include major restoration, endodontics, oral surgery, scaling and root planing, other periodontal treatment, prosthodontics, orthodontics, general anesthesia, other anesthesia, and adjunctive general (Table 1).

**Table 1:** Dental Procedure Categories.

| Group | Category | CDT Codes |
| --- | --- | --- |
| Preventive and Basic | Diagnostic | D0120-D0191 and D0414-D0999 |
|  | Imaging | D0210 – D0395 |
|  | Preventive | D1110 – D1999 |
| Major Dental Procedures | Minor Restoration | D2000 – D2664 |
|  | Major Restoration | D2710 – D2999 |
|  | Endodontics | D3000 -- D3999 |
|  | Oral Surgery | D7000 -- D7999 |
|  | Scaling & Root Planing | D4341 – D4355 |
|  | Other Periodontics | D4000 – D4340 and D4356 – D4999 |
|  | Prosthodontics & Implants | D5000 – D6999 |
|  | Orthodontics | D8000 – D8999 |
|  | General Anesthesia | D9220 – D9223 |
|  | Other Anesthesia | D9210-D9219 and D9230 – D9248 |
|  | Adjunctive General | D9110-D9120 and D9310 – D9999 |

Note: Categories make use of modified American Dental Association's Code on Dental Procedures and Nomenclature (CDT) 2018.

Medical spending on dental care includes dental services by a primary care provider, ambulatory surgeries for non-traumatic dental conditions (NTDC), emergency department (ED) visits for NTDC, and inpatient visits with NTDC. Dental conditions in the medical setting are defined as the range of ICD-10 diagnosis codes used in the Association of State & Territorial Dental Directors’ (ASTDD) definition of NTDCs.^31^ Spending on oral cancer is also included in this analysis and is defined as ICD-10 codes C00.0-C14.8 in either the inpatient or outpatient record. Finally, prescription drug costs for dental care or for medical spending on NTDC are included. Prescription drugs are defined as being prescribed for a dental condition if they occur within one day of an encounter with a dental procedure or an NTDC.

## Results

For Medicaid enrollees, spending on preventive and basic dental procedures increases from birth, peaking at age 7 at about $153 per year, reducing until age 11, with a second peak at age 14 at about $123 per year, and then falling dramatically, to a “valley” at age 22 at $16 per year. From this point, spending slowly increases through the early 40s age group and then decreases as people age (Figure 1a for spending by broad categories, Figure 1b for spending in detailed groupings, Supplemental Table 1, an interactive dashboard of the results is available at: https://public.tableau.com/profile/lifespan#!/). This pattern of spending on preventive and basic dental care is consistent with evidence on the burden of oral disease, especially early childhood caries, and reflects Medicaid policy by which states are required to provide dental coverage for children (generally up to age 20) but can choose to offer little or no dental coverage for adults.

**Figure 1a:**
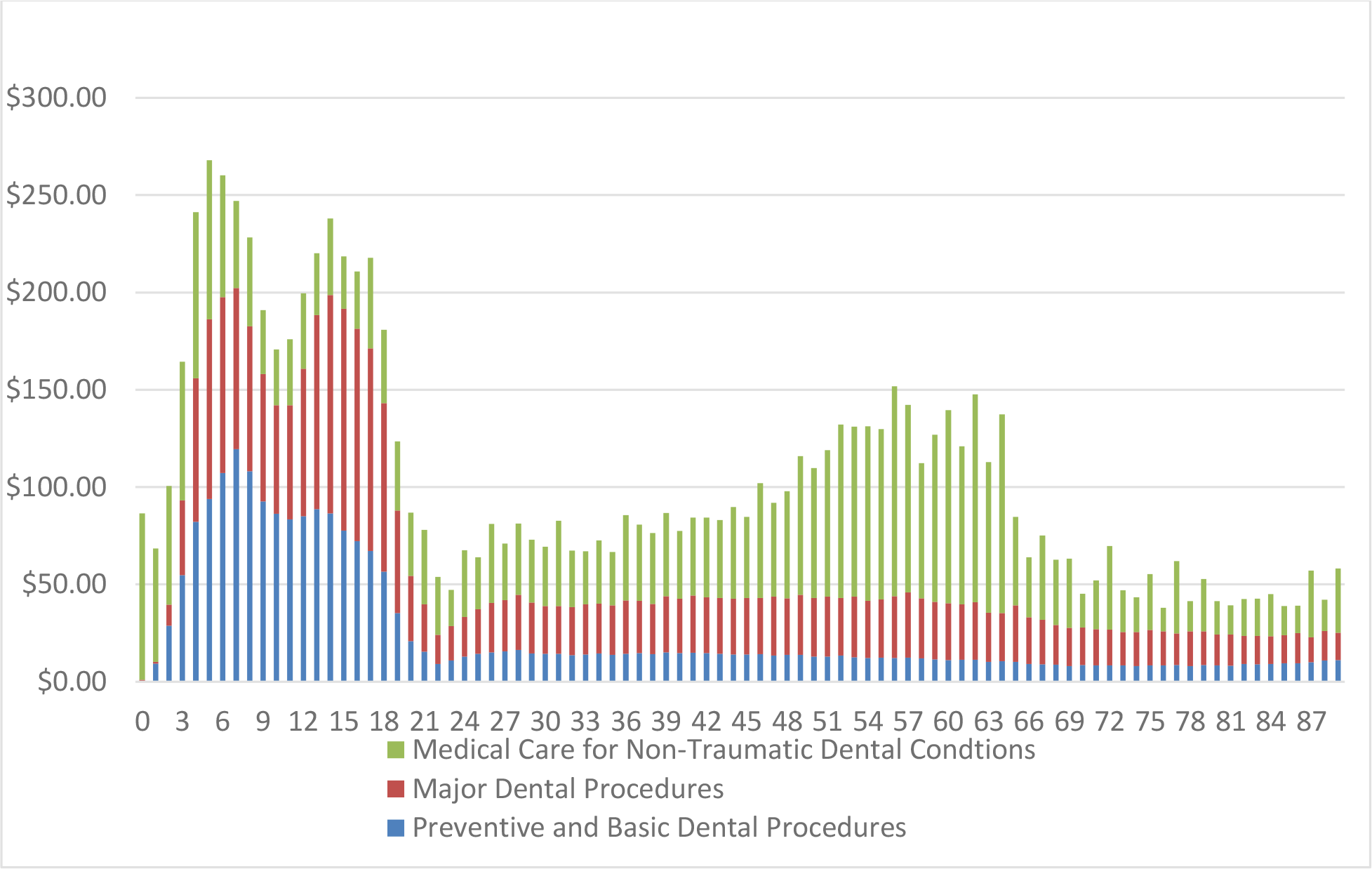
Average Per Enrollee Spending on Oral Health Care, in Broad Categories, Among Those Enrolled in Medicaid Over the Lifespan

**Figure 1b:**
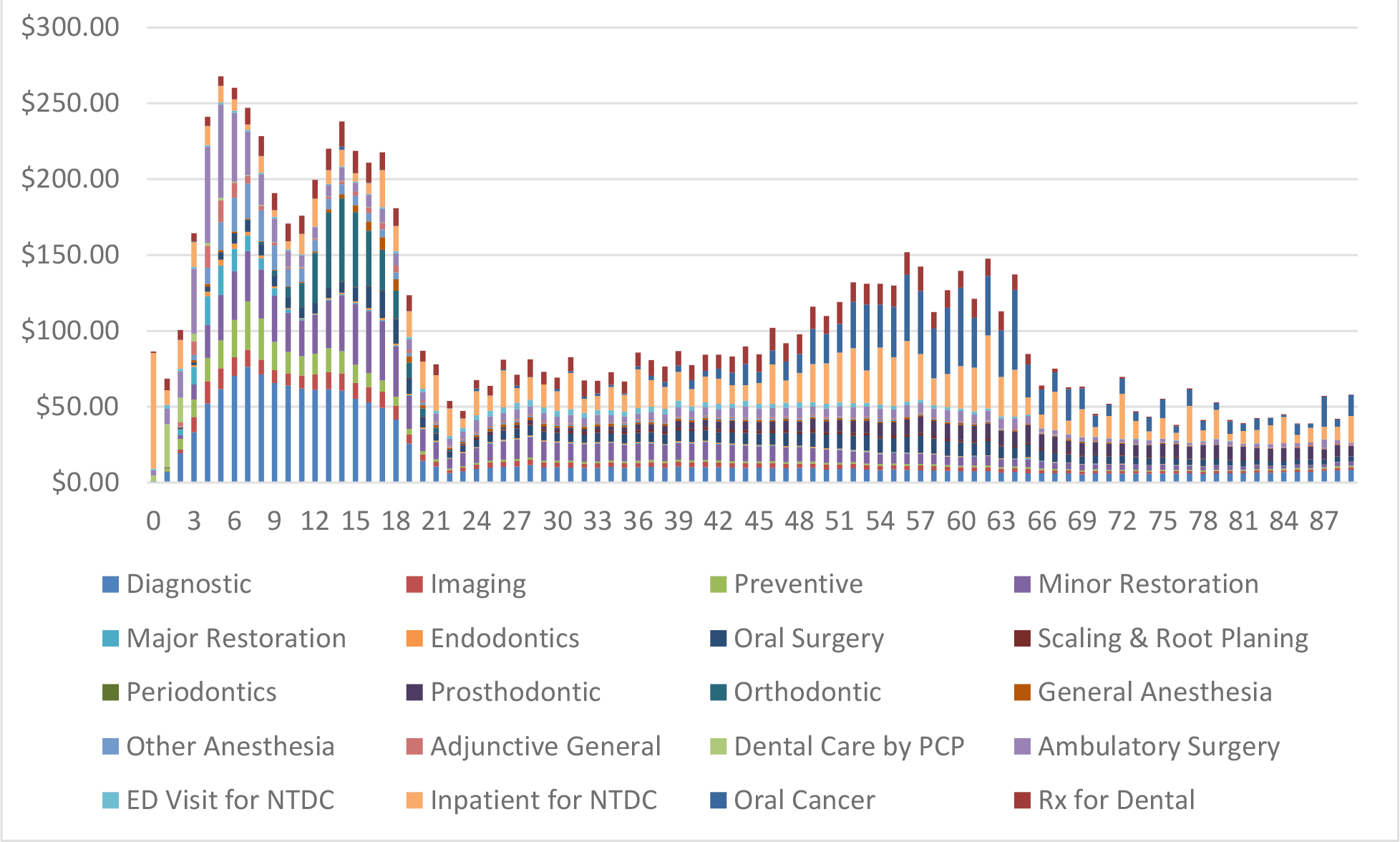
Average Per Enrollee Spending on Oral Health Care, in Detailed Categories, Among Those Enrolled in Medicaid Over the Lifespan

Spending on major dental procedures among those enrolled in Medicaid builds on the pattern seen in preventive dental care. Dental procedures associated with the surgical correction of early childhood caries, including major restoration and dental anesthesia, are prevalent among children under age 10 (Figure 1b). Spending on orthodontics spikes in adolescence, peaking at $54.62 per enrollee per year at age 14, and early adulthood. During this period, oral surgeries also become more common and make up the bulk of spending on major dental care procedures among older enrollees; spending on prosthodontics also increases for older adults, peaking at $11.51 per enrollee at age 65.

Among infants, most of the medical spending involves inpatient correction for oral health conditions shortly after birth (Figure 1b). Among children ages 1 and 2 enrolled in Medicaid, access to preventive and basic dental services is often provided by primary care providers (PCPs). Among 1-year olds, half of all oral health services are billed by a PCP, although reimbursement rates are higher so per enrollee spending is tilted towards PCPs (Supplemental Figures 3a and 3b). Between ages 3 and 10, ambulatory surgeries for NTDC are the dominant categories of medical spending on dental conditions, with spending peaking at age 4 at $63.52 per enrollee per year. ED visits make up a relatively small proportion of spending on dental care across all ages, but peaks among young adults in the oral health care valley described previously. Spending on oral cancer begins to increase by age 40 and peaks in the early 60s. Among adults, medical spending on dental conditions is most of the total spending on dental conditions (Figure 1a).

Average spending on basic and major dental care is higher at all ages among those with commercial insurance or supplemental Medicare plans than for those enrolled in Medicaid (Figure 2a for spending by broad categories, Figure 2b for spending in detailed groupings, Supplemental Table 2, an interactive dashboard of the results is available at: https://public.tableau.com/profile/lifespan#!/). The pattern of spending on preventive and basic dental care is similar to Medicaid, with spending increasing from birth, peaking at age 7 at about $265 per year, dropping again until age 11, with a second peak at 14 at about $230 per year; these expenses fall until a valley at 26 at $113 per year, with slow increases through 65 and decreasing after that. However, the peaks and valleys are much smaller in this population than among Medicaid enrollees, and there is less evidence among those enrolled in commercial insurance plans of a peak associated with early childhood caries.

**Figure 2a:**
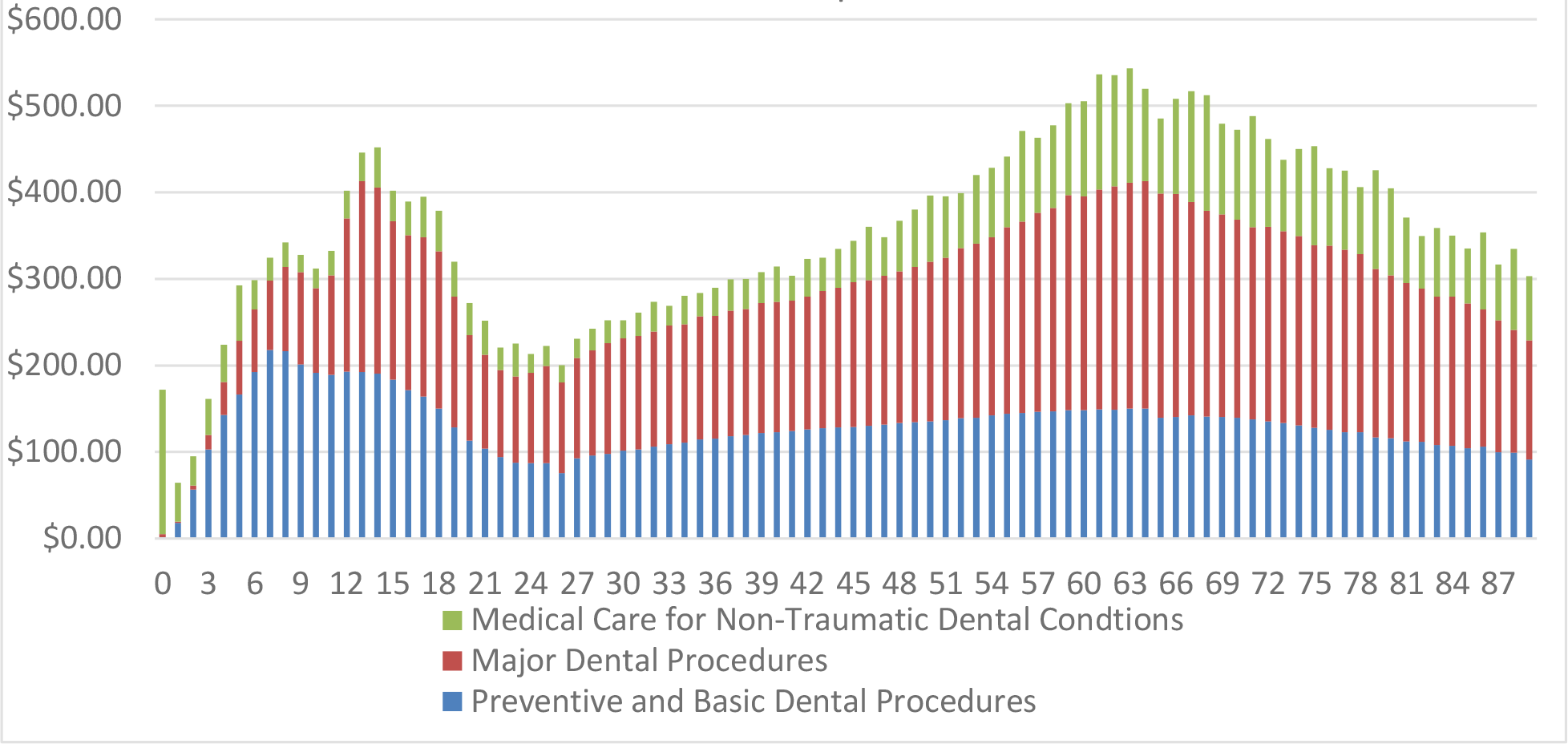
Average Per Enrollee Spending on Oral Health Care, in Broad Categories, Among Those Enrolled in Commerical Insurance Over the Lifespan

**Figure 2b:**
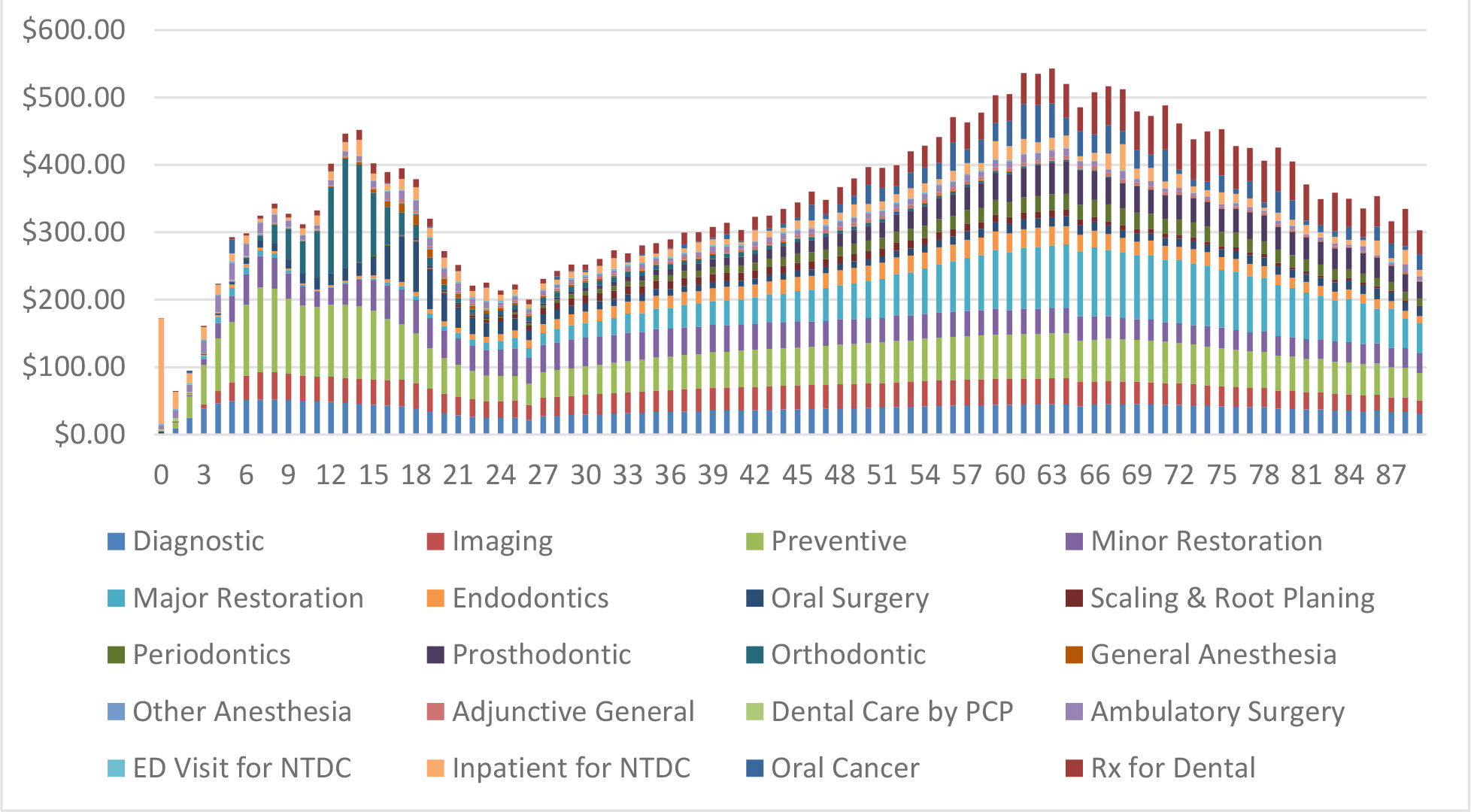
Average Per Enrollee Spending on Oral Health Care, in Detailed Categories, Among Those Enrolled in Commerical Insurance Over the Lifespan

Among those enrolled in commercial insurance plans, major dental care is far more common than in Medicaid populations, consistent with policy coverage differences. Until age 7, relatively little is spent on major dental care, never exceeding more than $33 per enrollee per year (Figure 2a). Between ages 9 and 16, spending on orthodontics becomes common, peaking at age 13 with $162 in average spending per year (Figure 2b). In late adolescence, oral surgeries become more common, peaking at age 18 at $75 per enrollee. Among adults, major restorations, such as crowns and endodontic procedures, dominate spending on dental care, along with prosthodontics. Average spending peaks at age 66 at $222 per enrollee and declines afterward.

Overall, medical spending on NTDC and oral cancer makes up a much smaller proportion of overall spending at each year of life in the commercially insured population than among those enrolled in Medicaid, except for infants (Figure 2a). Dental care by primary care providers and ambulatory surgeries comprises a very small proportion of overall spending within the commercially insured population, especially in relation to Medicaid populations; this likely reflects differences in reimbursement patterns and reduced emphasis on interprofessional practice in this setting (Figure 2b). Similarly, hospital visits for NTDC are relatively rare, although spending on prescription drugs for dental conditions is more common. Spending on oral cancer begins to increase by age 50 and peaks at 67.

Differences in per enrollee spending between the Medicaid and commercially insured populations partly reflect differences in utilization and reimbursement patterns. Utilization of dental care provided by dental offices is higher among those enrolled in commercial insurance for all ages (Supplemental Figure 3a). On the other hand, utilization of medical services for basic and preventive dental care, NTDCs and oral cancer is higher among every age group of Medicaid enrollees, except among those aged 65 or higher (Supplemental Figure 3b and 3c).

There are also notable differences with tooth mortality due to tooth extractions or removal. In supplemental analysis of the data, we find that individuals with Medicaid benefits are 5 times more likely to have a tooth extracted (CDT 7140, D7210, D7220, D7230, D7240, D7241) than those with commercial coverage, reflecting differences in reimbursement patterns and broader health inequities. Additionally, adults age 35 or older enrolled in Medicaid are 4.25 times more likely to have 6 or more teeth extracted in a single year compared to adults enrolled in commercial insurance plans. Tooth mortality is tightly tied to comorbidities and chronic disease, as the higher rates of diabetes, cardiovascular diseases, oral cancers, and substance abuse disorders in the Medicaid enrolled populations are all associated with increased tooth mortality.^32–35^

Average spending per enrollee on dental care is higher among those enrolled in commercial insurance plans, regardless of age (Supplemental Figure 4a). This pattern remains even with alternative methods of calculating costs. In fact, when costs are calculated using a usual and customary rate fee schedule, the difference is even more stark, especially among the period in which orthodontic care is common (Supplemental Figure 4b). However, reflecting the large differences in utilization and access to dental care due to policy variations, when averages are calculated per patient, the differences are less substantial among children and adults through their mid-40s, but spending does diverge among older adults (Supplemental Figure 4c).

## Discussion

This study represents one of the most comprehensive life course analyses of spending on dental care, utilizing data from over 31 million Medicaid claims and over 45 million commercial or Medicare supplemental claims. Combined, this data captures information from payment sources that cover 55% of the total dental spending in the United States (45% by commercial insurance and 10% by Medicaid), with the bulk of the rest (41%) being paid out of pocket while Medicare covers 1% and other insurers cover the remainder.^36^ Our analysis revealed several key findings. For both Medicaid and commercial structures, there is an overall deficiency in dental utilization and spending during the first 4 years of life (Figures 1a and 2a). Although both commercial and Medicaid systems see a reduction in utilization during the early 20s to mid-30s, Medicaid sees this decline continue throughout adulthood (Supplemental Figure 3a). Spending on oral cancer begins to increase in the early 40s and peaks in the early 60s with a total of $48 million spent in Medicaid and $69 million spent by commercial payers. Cumulatively, this investigation demonstrates unequal and inequitable access and outcomes of healthcare.

Previous studies also found that while overall oral health interactions are low during the first 4 years of life, medical teams play a significant role in oral health interactions for young children with 88% of all visits by age 1 and 52% of all visits up to age 4 in Medicaid and 70% of all visits by age one and 33% of all visits by age 4 in commercial insurance.^37–38^ Our analysis compliments this data illuminating the missed opportunity to engage young children in the recommended first dental visit by age one.

Because most low-income adults lack affordable access to preventive dental services, localized oral disease can begin and advance to more extensive and more acute conditions. This leaves these adults with few, if any, options other than visiting hospital EDs, a decision that may well be reflected by the higher rates of such visits among Medicaid-enrolled adults, starting in their 20s and continuing through their 50s, as seen in our analysis. This strongly suggests that many adults who lack the coverage to secure care eventually do become patients, albeit with oral conditions that are more acute and very costly to treat.

It is unclear the degree to which the observed oral cancer prevalence is shaped by the relatively low dental utilization rates among Medicaid enrolled adults. Better access to regular preventive dental care throughout adulthood years could have provided opportunities for education, anticipatory guidance on behaviors and risk factors, and earlier diagnosis. Additionally, the utilization of Human Papilloma Virus (HPV) vaccinations is still not widespread, limiting the ability to determine what impact its administration is having on overall incidence or prevalence.

While the current multi-payer structure allows for a diverse set of insurance products to beneficiaries and can create innovation, it also fortifies health disparities, discounts social determinants of health, and makes it difficult to achieve population health over the lifetime.^39–43^ The utilization and spending design of both commercial and Medicaid dental insurance is shaped by financial incentives that create business and care models influenced by revenue collection, risk pooling, purchasing, and the lack of social empathy.^44^ The cumulative effect produced a focus on finding or awaiting disease advancement as opposed to optimizing prevention and health, and rewarding lack of disease.

Some limitations should be considered when interpreting the results of this analysis. The first is that these are cross-sectional data based on 2018 claims only, which do not capture changes over time that may be evident through longitudinal analyses. Another weakness is that claims paid by Medicare are not included in the Medicaid database, making it possible that some claims are missing for those patients, especially those who are dual-eligible. Furthermore, Medicaid is administered at the state level, with significant variability in both pediatric and adult coverage (with relatively few states providing extensive adult oral health coverage), and the results presented may not be generalizable to all states and plans. Finally, people who pay 100% of their dental expenses out of pocket (the self-pay population), as well as those who may have received dental services at free clinics or charity care, are not present in the analysis

Future analysis should examine operating pathway variations through oral health and medical systems to demonstrate how inequalities in care at one point in time influence trajectories of care and systemic outcomes. A deeper understanding of variations of these patient pathways over a lifespan can allow researchers, payers, funders, and care teams to better understand how new care models, social programs, grant spending, and changes in political and socioeconomic environments impact society or quality of life.

## Conclusion

Dental coverage is one area of policy that can be informed by lifespan data. This unique lifespan analysis of the U.S. multi-payer dental care system demonstrates the complexities of the current dental service environment and conveys the difficulties patients can have maneuvering in a fragmented and siloed oral health system over their lifetime. Specifically, income inequality and the absence of social empathy provides more readily available care and a wider range of care options to individuals who happen to have a more extensive benefit plan and/or a better ability to pay out-of-pocket expenses. Regardless of the differences in equity, both private and public payment methodologies lack a prevention-first focus and include financial incentives that incentivize significant restorative care. Health policies should be focused on optimizing care delivery to provide effective preventive care at specific stages of the lifespan, as opposed to policy development that is precipitated by short-term political and financial dynamics, thus avoiding the necessity for major surgical dental care.^45^ Lifespan data could also positively inform programmatic strategies, such as medical-dental integration, and clinical practices, such as disease prevention and management.

## Data Availability

The data is availability through IBM Watson through a data use agreement.

### Appendices

**Appendix 1.**
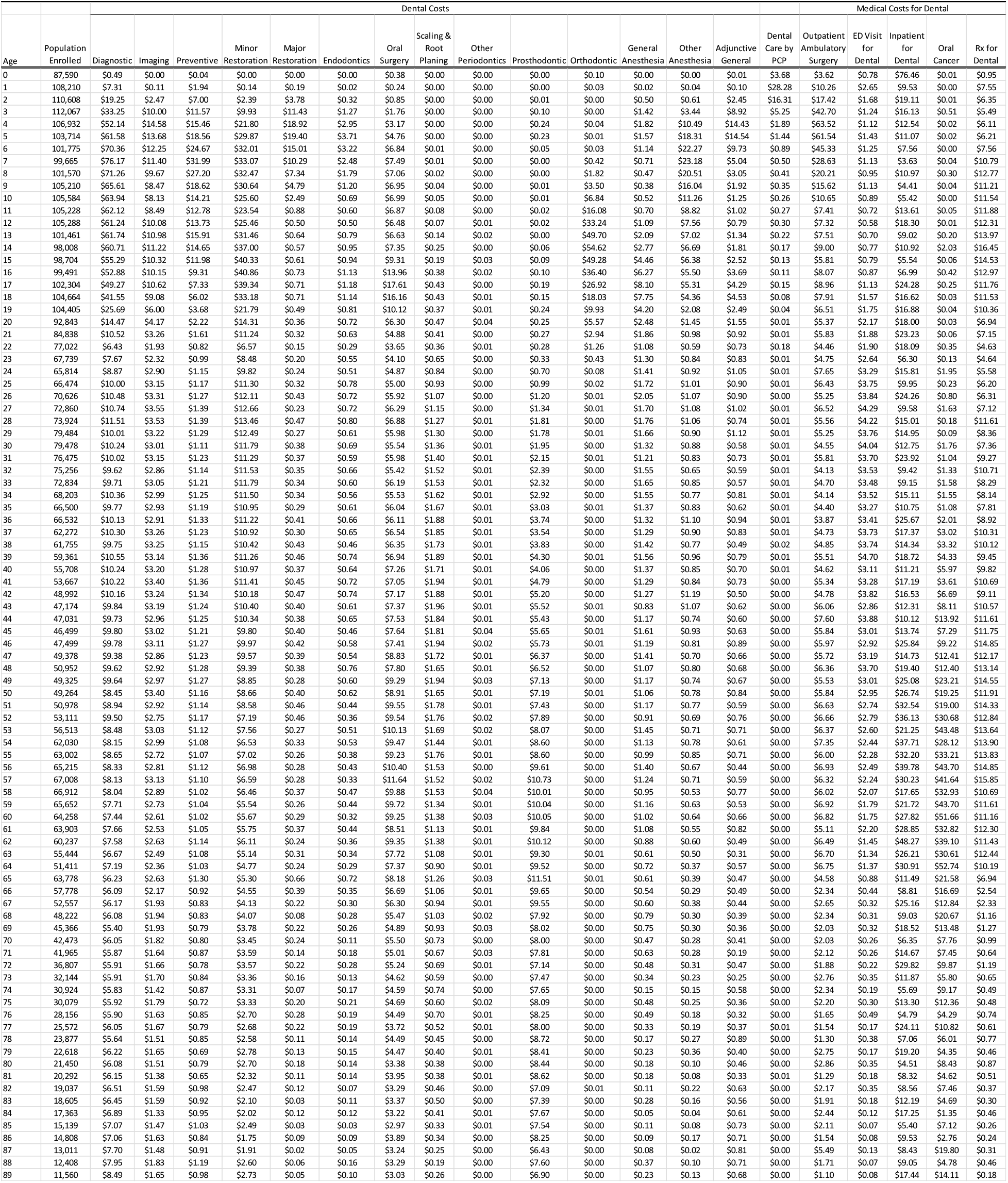

**Appendix 2.**
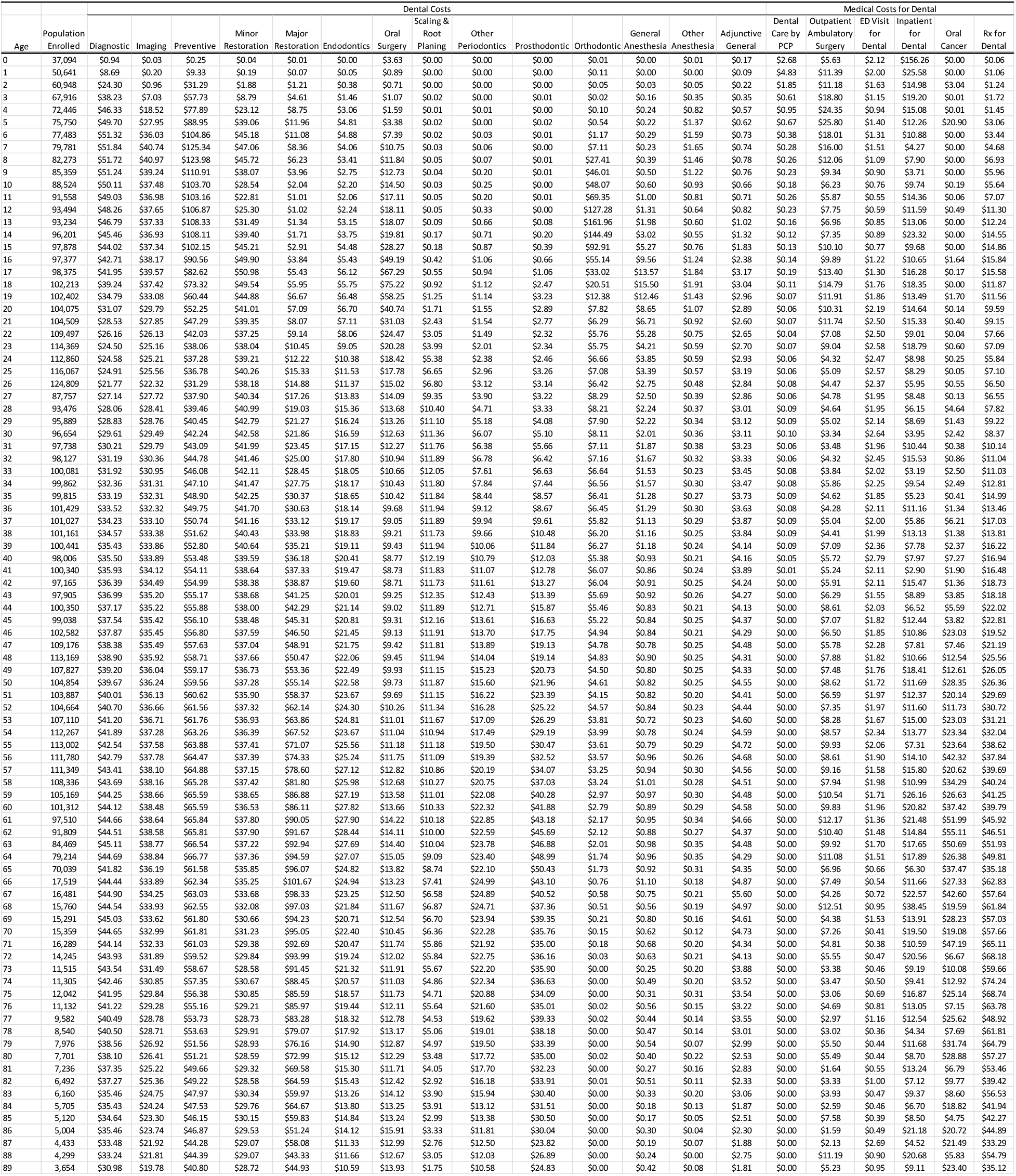

## Supplementary Material

**Supplemental Figure 3a:**
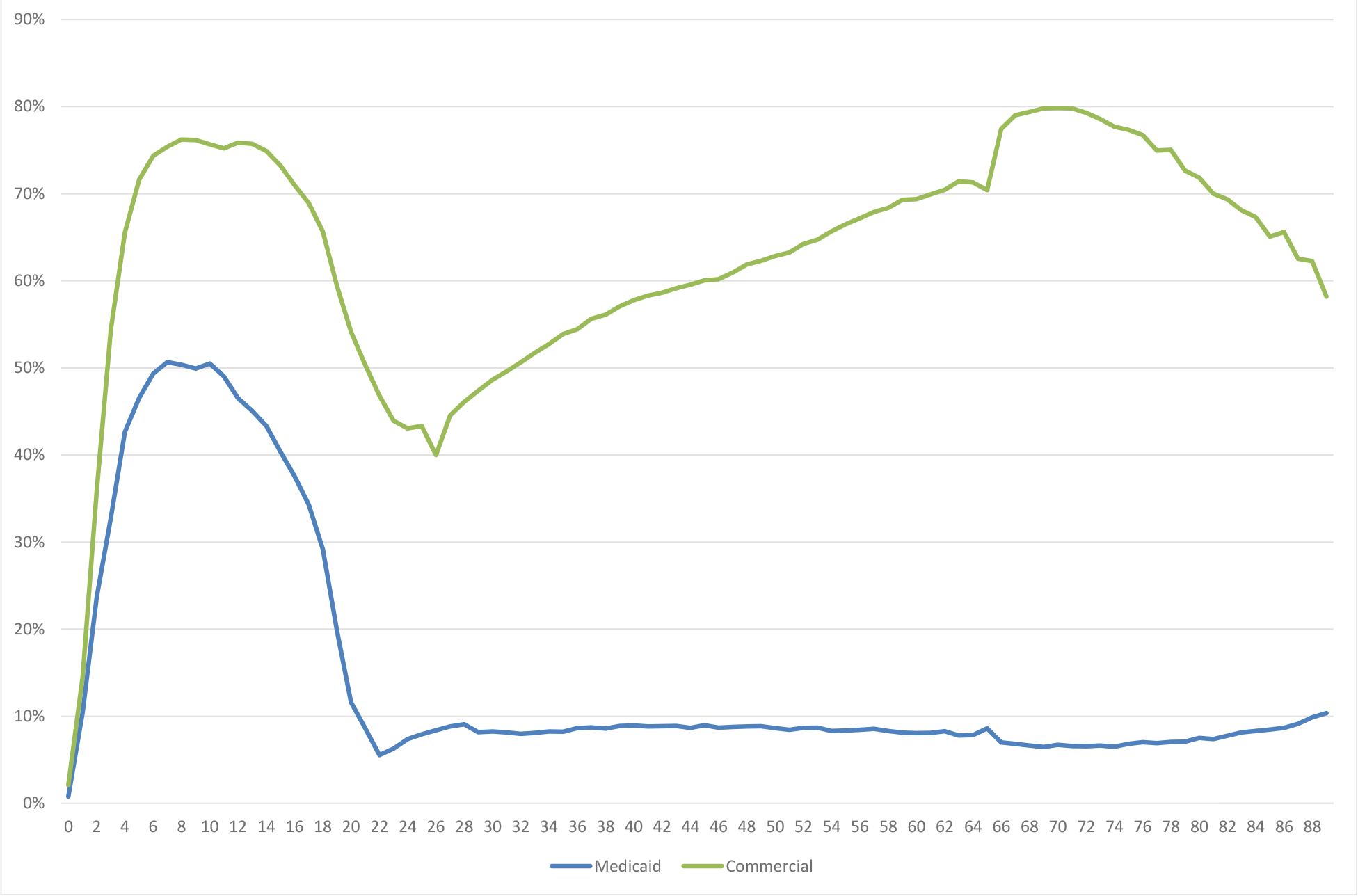
Utilization of Dental Care over the Lifespan among Medicaid and Commercially Insured Populations in 2018

**Supplemental Figure 3b:**
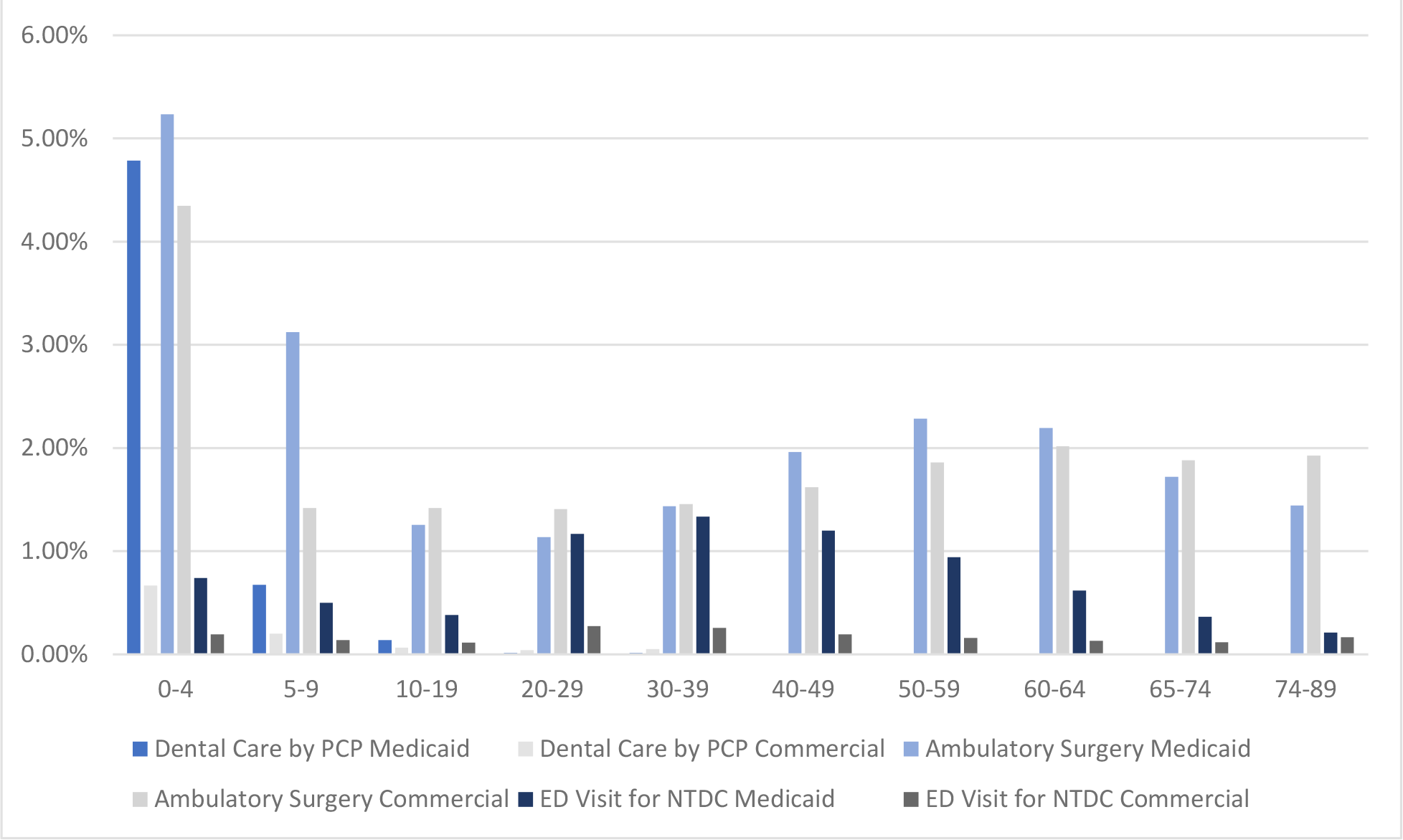
Utilization of Hospital Outpatient and ED for Dental Care among Medicaid and Commercial Insured Populations in 2018

**Supplemental Figure 3c:**
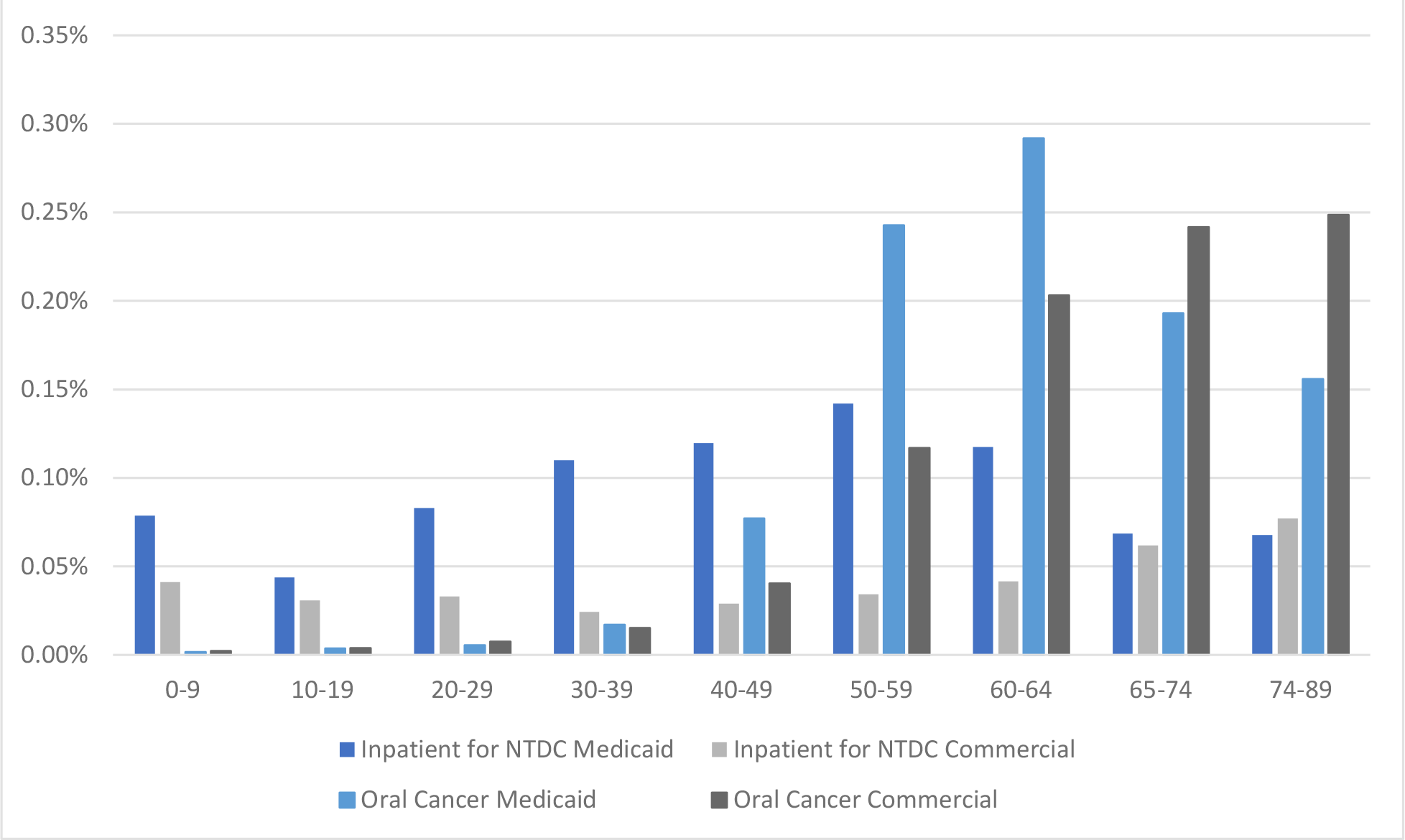
Utilization of Hospital Inpatient and Incidence of Oral Cancer among Medicaid and Commercial Insured Populations in 2018

**Supplemental Figure 4a:**
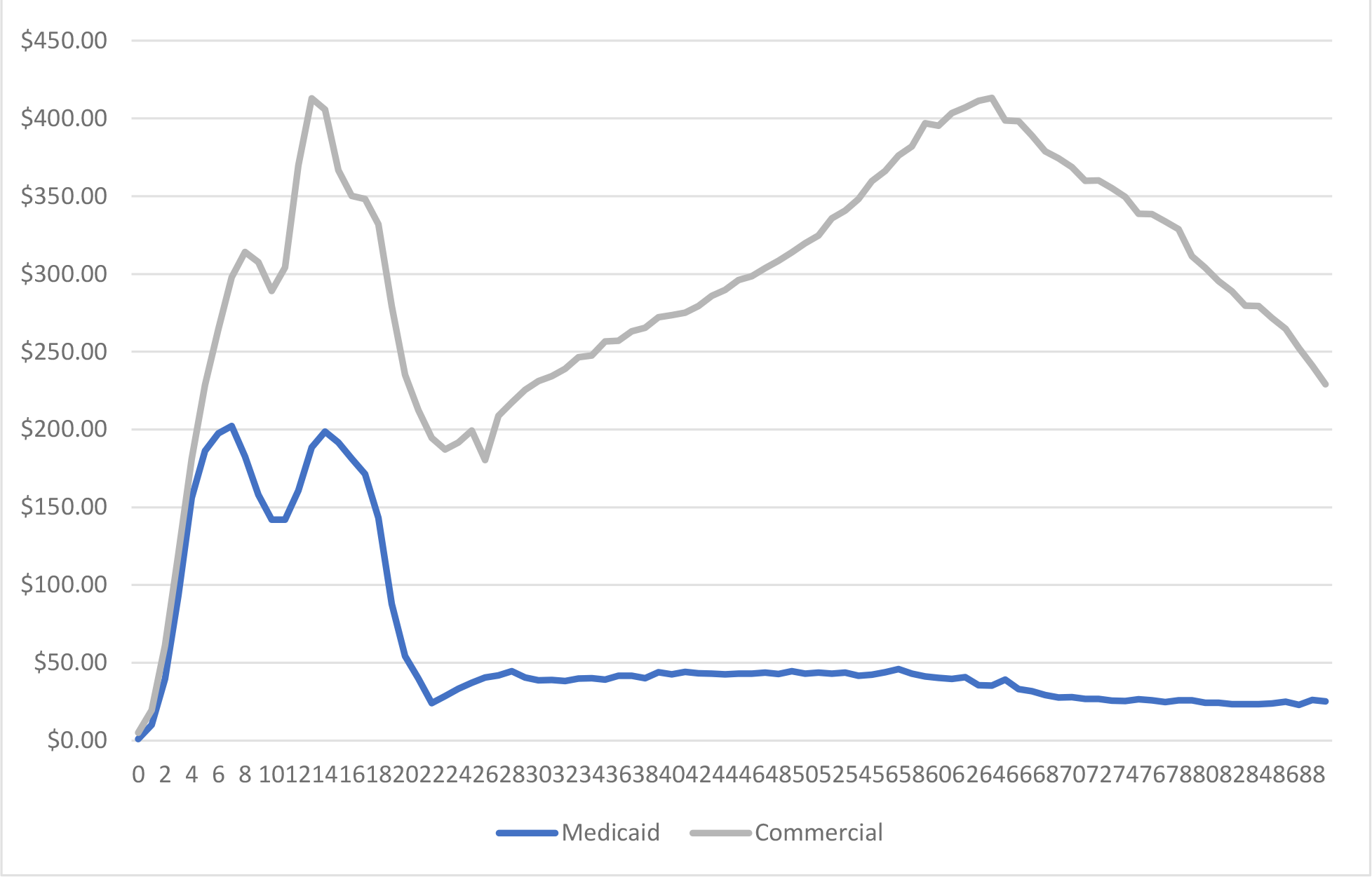
Average Per Enrollee Payment to Dental Provider

**Supplemental Figure 4b:**
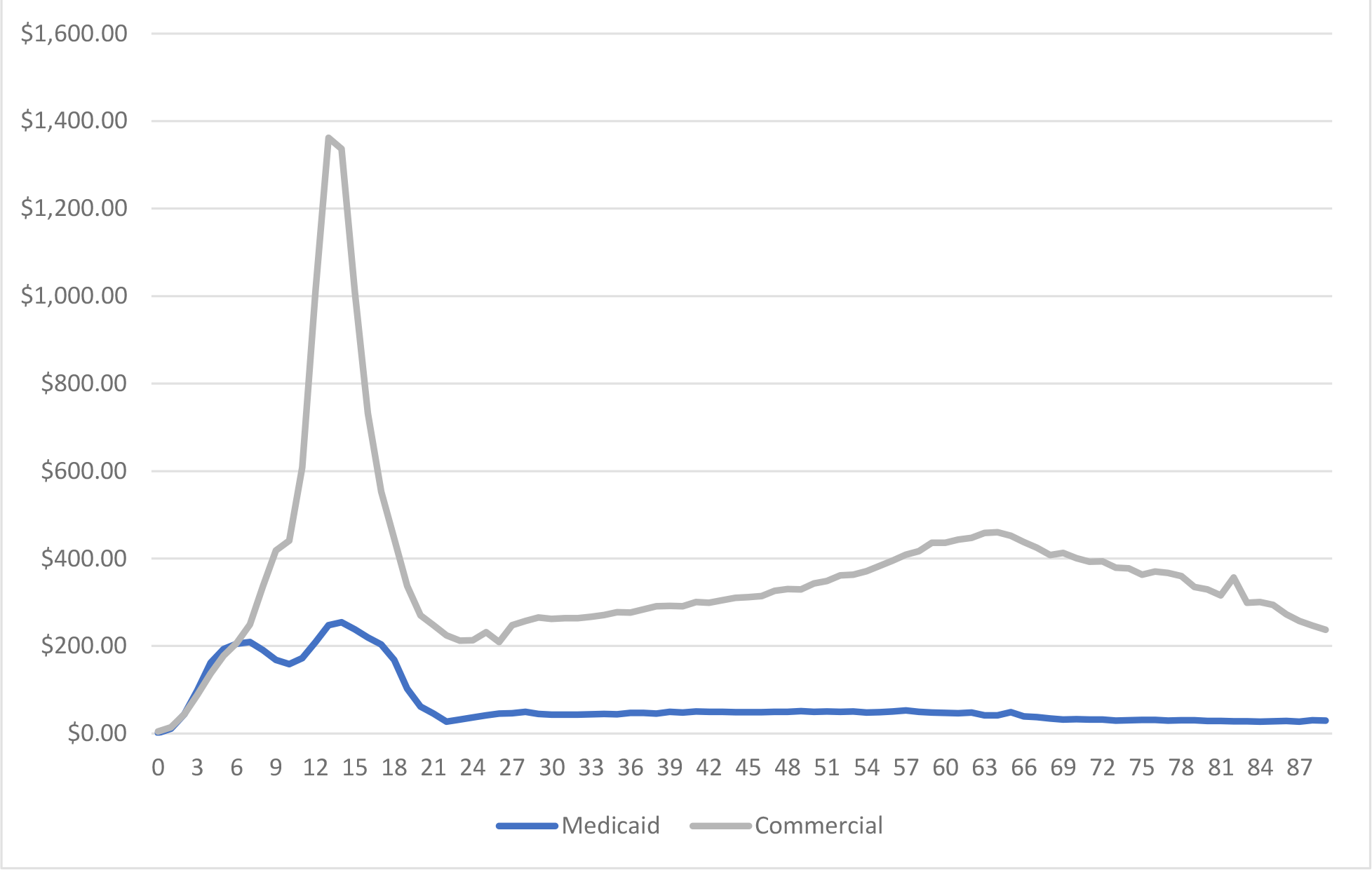
Average Per Enrollee Cost Using UCR Fee Schedule

**Supplemental Figure 4c:**
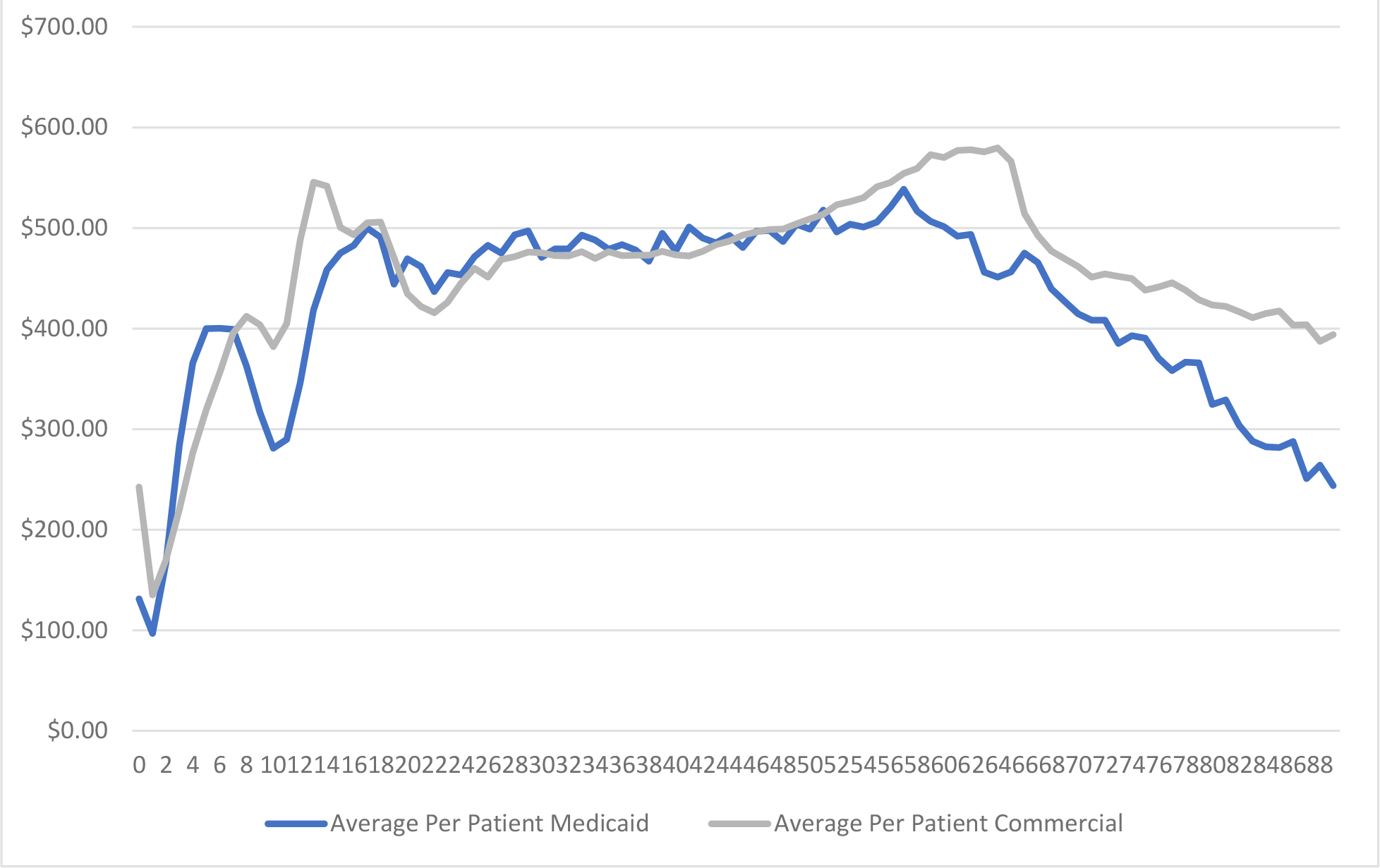
Average Per Patient Payment to Dental Provider

